# Barriers and facilitators to intracerebral haemorrhage platform trial recruitment: a survey of stroke clinicians

**DOI:** 10.64898/2026.03.05.26347732

**Authors:** Anuka Boldbaatar, Tom J Moullaali, Allan MacRaild, Sarah Risbridger, Alice Hosking, Connor Richardson, Gwynneth A Clay, Martin Dennis, Nikola Sprigg, Mark Barber, Adrian R Parry-Jones, Christopher J Weir, David J Werring, Rustam Al-Shahi Salman, Neshika Samarasekera

**Author notes:** **Corresponding author:** Centre for Clinical Brain Sciences, The University of Edinburgh, Edinburgh, EH16 4SB, UK.

## Abstract

**Background:** Platform trials are an efficient trial design which enable testing of multiple interventions simultaneously. They could advance knowledge of treatments for intracerebral haemorrhage (ICH). We aimed to investigate the views of clinicians involved in stroke research on recruitment to a future platform trial for ICH.

**Methods:** Between April and July 2025, we conducted a UK-wide online survey of clinicians actively involved in stroke research using convenience sampling through professional organisations. Participants considered factors related to the consent process and research environment and could provide optional free text responses about additional barriers or facilitators to recruitment. We used descriptive statistics for quantitative data and content analysis for qualitative data.

**Results:** Among 73 respondents, 46 (63%) were female, 36 (50%) were stroke physicians, 24 (34%) nurses, 6 (8%) allied health professionals, and 7 (10%) were in other roles. 36 (49%) had >20 years’ clinical experience, 45 (61%) reported spending <10% of their role in research. 66 (91%) thought that a platform trial would be a good option for testing interventions for patients with stroke due to ICH. Across 11 modifiable factors, clinicians most frequently rated perceived importance of the research question as a facilitator of recruitment (94%), while clinician preference for specific treatments was most frequently rated as a barrier (48%). Two themes emerged from free text responses: study design and infrastructure. Regarding study design respondents perceived consent procedures (n=9), study materials (n=8), study procedures (n=8), eligibility assessment (n=6), the research question (n=3) and randomization (n=3) as important for a future platform trial. Regarding infrastructure, emergent factors were staffing (n=17), local research culture and capacity (n=9), research governance and delivery (n=6), and training (n=6).

**Conclusion:** The overwhelming majority of respondents from the UK clinical stroke community supported a platform trial for ICH, although the influence of survey responder bias is unknown.

## Introduction

Despite advances in the acute management of intracerebral haemorrhage (ICH) through integrated bundles of care,^1^ we still do not have a specific medical treatment for ICH. Randomised controlled trials of interventions for ICH have been small and are often time-consuming to conduct.^2^ Platform trials allow testing of multiple interventions simultaneously and are an efficient trial design which use a shared infrastructure, so have the potential to hasten the identification of effective medical treatments.^3^ When setting up a platform trial, involvement of relevant stakeholders early is essential to facilitate engagement and may inform aspects including trial design.^4^ Therefore, we used a cross-sectional online survey to evaluate the views of clinicians involved in stroke research about recruitment to a future platform trial for patients with stroke due to ICH.

## Methods

### Study Design and Participant Recruitment

We followed Consensus-Based Checklist for Reporting of Survey Studies (CROSS)^5^ for reporting.

We developed the survey (supplement) by drawing on relevant literature on barriers and facilitators to recruitment in randomised controlled trials (RCTs)^6,7^ and clinical experience. We collected information about demographics, employment, and views on platform trials. Participants rated 11 potentially modifiable factors about the consent process and the research environment which could affect recruitment, using a 5-point Likert scale ranging from “strong barrier” to “strong facilitator”. Participants could also provide free text responses about other modifiable barriers and facilitators to recruitment. We did not undertake a formal sample size calculation as this was an open, convenience sample survey.

Clinicians (allied health professionals, nurses, and physicians) actively involved in stroke research were eligible to take part. We obtained written informed consent from all participants. We obtained ethical approval from the University of Edinburgh Research Ethics Committee (25-EMREC-009). We collected data anonymously and stored and handled it securely, according to institutional guidelines.

### Data collection

We used convenience sampling via professional organisations. We sent an email invitation to distribute the survey through the National Institute for Health and Care Research (NIHR) Stroke Research delivery network (United Kingdom), UK-wide National Stroke Nurse Forum, British and Irish Association of Stroke Physicians, Association of British Neurologists, Scotland-wide distribution list for stroke education sessions and NHS (National Health Service) Research Scotland Stroke research network between April 17, 2025, and July 17, 2025 with one additional reminder during the data collection period.

### Data analysis

We summarised participant demographics, employment characteristics, and survey responses using descriptive statistics. We conducted the analyses in Microsoft Excel (version 2512) and excluded missing values. We analyzed free text responses using qualitative content analysis.^8^ AB read the free text responses in full and performed inductive coding. A coding framework was developed comprising two overarching categories and associated subcategories to capture recurring issues across responses. Responses were coded manually given the modest volume of free text data, with responses permitted to be assigned to more than one subcategory where relevant. NS independently reviewed the coding framework and coded output, and the final category structure was agreed through discussion. We summarised the number of responses coded to each subcategory.

## Results

A total of 73 clinicians completed the survey. 46 (63%) were female, 32 (44%) were aged 40–49 years and 40 (55%) were based in England with the remainder based in Scotland (supplement, table 1). 36 (50%) respondents were stroke physicians, 12 (17%) were stroke nurse practitioners / specialist nurses, 12 (17%) were research nurses, 6 (8%) were allied health professionals, and 7 (10%) were in other roles. 36 (49%) had >20 years’ clinical experience and 45 (61%) reported spending <10 % of their role in research (supplement table 1).

All participants rated factors affecting platform trial recruitment. Of 73 respondents, 30 (41%) provided free text comments on potential modifiable facilitators, and 28 (38%) provided suggestions regarding modifiable barriers to recruitment.

### Clinicians’ views on platform trial feasibility

66 of 73 (91%) respondents thought that a platform trial would be a good option for testing interventions for patients with stroke due to ICH.

### Perceived Barriers and Facilitators to Recruitment

59 (82%) respondents viewed provision of brief, user-friendly participant information as a facilitator and 58 (79%) valued the option to share it electronically with participants. 61 (84%) felt that the ability to seek verbal consent before written consent in hyperacute settings was a facilitator (figure 1).

**Figure 1.**
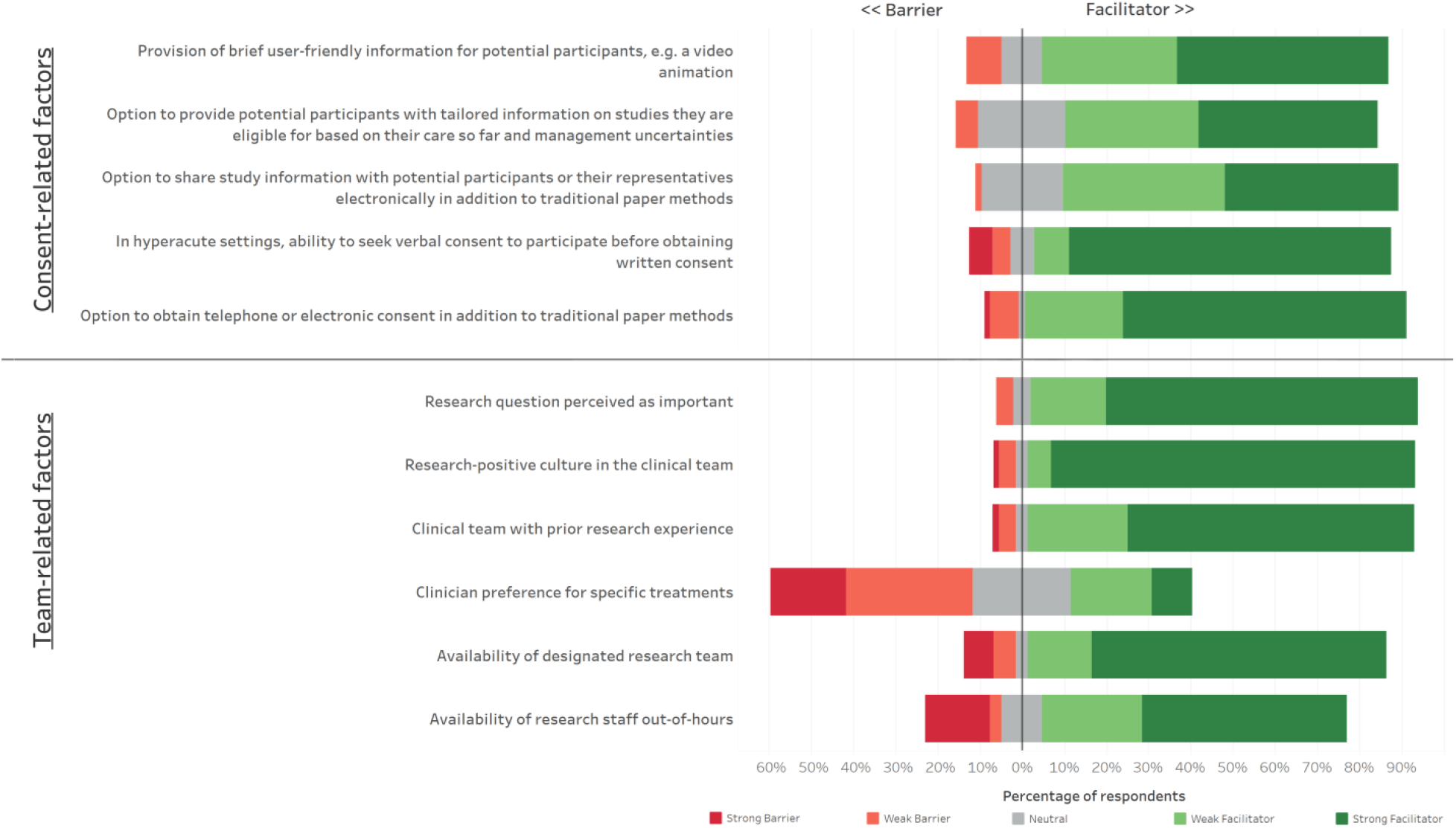
Divergent bar chart of participant ratings to the consent-related and team-related factors centred on the neutral response

67 (91%) perceived having a research culture within the clinical team as a facilitator and 66 (92%) viewed prior research experience as a facilitator. 35 (48%) thought that clinician preference for a particular treatment was a barrier.

### Qualitative content analysis

The first 50 survey respondents provided 20 free text responses about potential facilitators and 20 responses about potential barriers to recruitment (supplement). From these we derived two overarching categories, study design and research infrastructure, and we did not derive any new categories from the remaining 23 respondents who provided 18 free text responses (supplement). Two responses described participant-level barriers that did not map to either of the two main categories.

Responses relating to study design centred on making trial procedures as streamlined and pragmatic as possible to minimise burden on both staff and participants. The most frequently cited study design factor related to consent processes (n=9). In addition to streamlined and inclusive consent processes, some responses outlined access to a professional legal representative (an independent clinician) and deferred (retrospective) consent as key approaches in emergency settings. Eight respondents emphasised the value of clear, accessible study materials tailored both to patients and the recruitment setting. Suggested solutions included using simplified language, providing materials in different languages and incorporating visual formats such as videos. Other responses described study procedures (n=8), identifying eligible patients (n=6), the research question (n=3) and randomization (n=3). Figure 2 summarizes emergent study design factors, mapped across key stages of ICH platform trial design.

**Figure 2.**
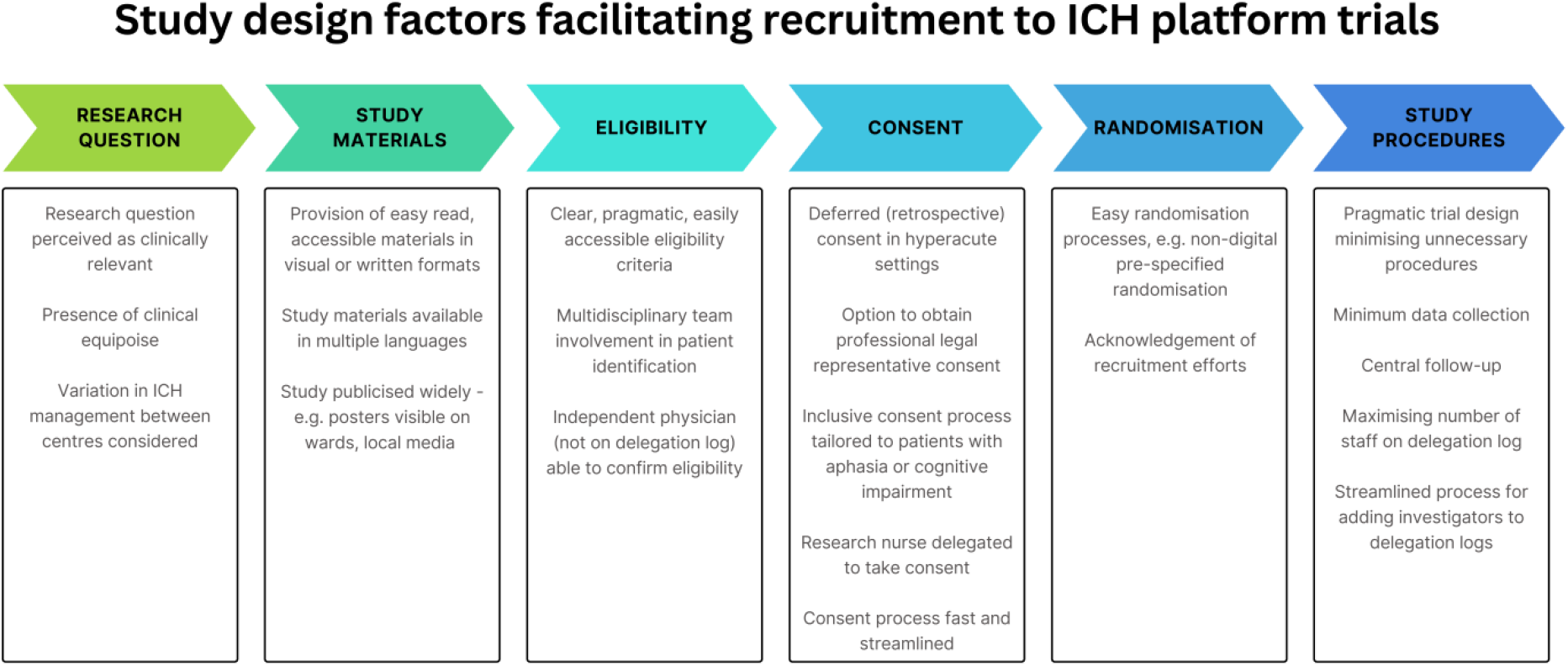
Study design factors facilitating recruitment to ICH platform trials.

### Infrastructure-related factors

Most responses about research infrastructure concerned staffing (n=17). Respondents emphasised the importance of adequate full-time research staffing to support multi-site delivery and enable out-of-hours recruitment. They noted the value of broadening research opportunities across the multidisciplinary team and protecting research activity in clinical job plans, giving staff protected time and formal responsibility for screening and recruitment. With support, respondents noted that out-of-hours recruitment was feasible without reliance solely on dedicated research staff.

Nine respondents noted factors pertaining to local research culture and capacity. Respondents perceived a research-positive culture as important and described constraints related to embedding research activity within clinical teams and routine workflows.

Respondents described research governance and delivery factors operating at both local and national levels (n=6). These included governance processes and associated timelines, administrative burden, and nationally coordinated ICH research delivery. Respondents perceived digital integration of trial infrastructure, for example through the NHS App, as a potential facilitator of recruitment.

Six respondents identified training as a facilitator, citing education on platform trial designs, refresher training, and support for clinicians less familiar with adaptive and platform methodologies.

## Discussion

In this survey of UK stroke clinicians, 91% of respondents thought a platform trial was a good option for testing interventions for ICH. Respondents identified factors related to study design which could enhance recruitment, including accessible study materials, an important research question and streamlined consent processes with the ability to seek verbal consent before written consent in hyperacute settings. Respondents also identified team, local site, and system-level factors that could influence recruitment, including research-positive culture, staffing capacity and training.

To our knowledge, this is the first survey to explore stroke clinicians’ views on platform trial recruitment, and most respondents supported a platform trial for ICH. Our findings are in keeping with the views of staff in the paediatric intensive care unit setting who were surveyed about developing a platform trial and thought it was ‘acceptable’ and a resource and cost-effective way to do clinical trials.^9^

Respondents affirmed the need for streamlined consent processes in time-critical settings including consideration of deferred consent.^10^ Hyperacute stroke trials have used different methods including deferred consent,^10^ verbal consent before written consent^11^ and initial brief written consent followed by full consent after randomisation.^12^ Although patients disagreed with deferred consent in the ESCAPE trial,^10^ this may be because participants in ESCAPE might have been randomised to the control (medical management) arm of the study rather than the intervention (thrombectomy) for which there was some evidence of efficacy at the time. Deferred consent was acceptable to 95% participants in the Alteplase Compared to Tenecteplase (AcT) trial.^11^ In both paediatric and stroke settings, patients/caregivers taking part in focus groups accepted that deferred consent could facilitate timely randomization in a platform trial.^9,13^ Following a public consultation, UK clinical trials regulatory reforms^14^ include a planned provision to allow simplified arrangements for seeking and evidencing consent in low-risk trials.

In keeping with other studies,^15,16^ respondents highlighted the need for study information to be brief and accessible, for example using user-friendly formats such as video or animation. They supported digital sharing of study information to enable access for families who may not be present on site. Studies of trial recruitment for other conditions, such as cancer, suggest that recruitment is not facilitated solely by accessibility, but also by how much information resonates with patients’ priorities and concerns.^17^ Therefore, delivery methods which support flexibility and personalisation may help manage the informational demands of platform trials,^18^ within the practical constraints of recruitment in acute stroke care.

Our findings indicate that clinicians’ engagement with recruitment may be shaped by how meaningful they perceive the research question to be. Where perceived clinical value is low, clinicians may act as informal gatekeepers, selectively introducing research opportunities.^19^ Strong preferences for specific treatments were identified as a barrier. This is pertinent in ICH, where both between-site pathway variation^20^ and between-clinician differences^21^ may influence how uncertainty is framed when explaining the study. Practice variation should be addressed at the design stage by specifying core recruitment messages, defining the usual care context and eligibility pathways, and planning consistent recruitment communication as study and practice evolve.

Respondents stated the importance of training about platform trials. This is consistent with evidence that recruiters’ knowledge and confidence can influence participation decisions,^22^ and that limited understanding of research methodology can act as a barrier to recruitment.^23^ Since platform trials remain relatively unfamiliar to many clinicians and place additional demands on their ability to explain the study,^18^ team-wide research experience and training may be particularly important.

Respondents emphasised minimising the practical burden of trial delivery including protocol complexity, administration, and operational demands across eligibility assessment, randomisation, consent and study procedures. In platform trials, these challenges may be amplified by comparison-specific eligibility criteria^3^ and increasingly complex or fragmented databases.^24^ Together, these findings indicate that platform trials should prioritise operational simplicity and feasibility, as delivery burden is likely to influence sustained clinician engagement with recruitment.

This study has several strengths. Respondents had a range of professional roles, providing breadth of clinical and research experience. Most respondents reported clinical roles, suggesting the views captured are broadly representative of clinicians directly involved in recruitment. We also captured valuable qualitative insights from free text responses.

This study has limitations. As dissemination occurred via multiple networks with overlapping membership, the number of eligible recipients could not be determined so we are unable to calculate a response rate. We cannot exclude duplicate responses due to an open survey link, although the risk is likely low given the professional nature of the respondent group and absence of incentives. Clinicians from Wales and Northern Ireland were not represented, which may limit generalisability. Findings may be subject to responder bias if clinicians with more favourable views of platform trials were more likely to respond and, as a UK survey, it may not apply to settings with different research governance, resourcing, and trial delivery infrastructure.

In this survey of UK stroke clinicians, 91% of respondents thought that a platform trial would be good option for testing interventions for ICH. Future work should test and refine proportionate consent approaches for ICH platform trials, including staged consent pathways. Qualitative work with patients and families affected by ICH would complement clinician perspectives and help ensure recruitment approaches are acceptable, equitable, and ethically robust.

## Data Availability

All data produced in the present study are available upon reasonable request to the authors

