## Supplemental Material for "Barriers and facilitators to intracerebral haemorrhage platform trial recruitment: a survey of stroke clinicians"

#### Additional survey methods

The survey was hosted on Jisc Online Surveys.[25] As recruitment was supported by onward sharing, respondent-level access control was not feasible, and duplicate submissions could not be technically prevented.

**Table S1 Participant characteristics**

| | n[a] | % |
|---|---|---|
| **Professional role** | | |
| Stroke Physician | 36 | 49% |
| Stroke Nurse Practitioner / Specialist Nurse | 12 | 16% |
| Research Nurse | 12 | 16% |
| Other Physician | 6 | 8% |
| Allied Health Professional | 6 | 8% |
| Clinical Psychologist | 1 | 1% |
| **Proportion of role spent in research** | | |
| <10% | 44 | 61% |
| >50% | 15 | 21% |
| 10-25% | 8 | 11% |
| 25-50% | 6 | 8% |
| **Years of experience** | | |
| <5 years | 6 | 8% |
| 5-10 years | 10 | 14% |
| 10-20 years | 22 | 31% |
| 20+ years | 35 | 49% |
| **Region** | | |
| England | 40 | 55% |
| Scotland | 33 | 45% |
| **Age** | | |
| <30 | 4 | 5% |
| 30-39 | 11 | 15% |
| 40-49 | 32 | 44% |
| 50-59 | 21 | 29% |
| 60+ | 5 | 7% |
| **Gender** | | |
| Female | 46 | 63% |

| | | |
|---|---|---|
| Male | 26 | 36% |
| Prefer not to say | 1 | 1% |

a) Total n = 73. There were no missing data for any demographic variables presented

**Table S2 Distribution of clinician responses to 11 factors affecting recruitment to ICH platform trials (n=73)**

| Factors affecting recruitment to ICH platform trials | Strong Barrier | Weak Barrier | Neutral | Weak Facilitator | Strong Facilitator |
|---|---|---|---|---|---|
| Provision of brief user-friendly information for potential participants, e.g. a video animation* | 0 (0) | 6 (8) | 7 (10) | 23 (32) | 36 (50) |
| Option to provide potential participants with tailored information on studies they are eligible for based on their care so far and management uncertainties | 0 (0) | 4 (5) | 15 (21) | 23 (32) | 31 (42) |
| Option to share study information with potential participants or their representatives electronically in addition to traditional paper methods | 0 (0) | 1 (1) | 14 (19) | 28 (38) | 30 (41) |
| In hyperacute settings, ability to seek verbal consent to participate before obtaining written consent** | 4 (6) | 3 (4) | 4 (6) | 5 (8) | 55 (76) |
| Option to obtain telephone or electronic consent in addition to traditional paper methods | 1 (1) | 5 (7) | 1 (1) | 17 (23) | 49 (67) |
| Research question perceived as important | 0 (0) | 3 (4) | 3 (4) | 13 (18) | 54 (74) |
| Research-positive culture in the clinical team | 1 (1) | 3 (4) | 2 (3) | 4 (5) | 63 (86) |
| Clinical team with prior research experience* | 1 (1) | 3 (4) | 2 (3) | 17 (24) | 49 (68) |
| Clinician preference for specific treatments | 13 (18) | 22 (30) | 17 (23) | 14 (19) | 7 (10) |
| Availability of designated research team | 5 (7) | 4 (5) | 2 (3) | 11 (15) | 51 (70) |
| Availability of research staff out-of-hours* | 11 (15) | 2 (3) | 7 (10) | 17 (24) | 35 (49) |

Values are count (percentage) of respondents per item. *One missing value. **Two missing values

### Checklist for Reporting Of Survey Studies (CROSS)[5]

| Section/topic | Item | Item description | Reported on page # |
|---|---|---|---|
| **Title and abstract** | | | |
| Title and abstract | 1a | State the word "survey" along with a commonly used term in title or abstract to introduce the study's design. | 1 |
| | 1b | Provide an informative summary in the abstract, covering background, objectives, methods, findings/results, interpretation/discussion, and conclusions. | 2 |
| **Introduction** | | | |
| Background | 2 | Provide a background about the rationale of study, what has been previously done, and why this survey is needed. | 3 |
| Purpose/aim | 3 | Identify specific purposes, aims, goals, or objectives of the study. | 3 |
| **Methods** | | | |
| Study design | 4 | Specify the study design in the methods section with a commonly used term (e.g., cross-sectional or longitudinal). | 3 |
| Data collection methods | 5a | Describe the questionnaire (e.g., number of sections, number of questions, number and names of instruments used). | 3 |
| | 5b | Describe all questionnaire instruments that were used in the survey to measure particular concepts. Report target population, reported validity and reliability information, scoring/classification procedure, and reference links (if any). | 3 |
| | 5c | Provide information on pretesting of the questionnaire, if performed (in the article or in an online supplement). Report the method of pretesting, number of times questionnaire was pre-tested, number and demographics of participants used for pretesting, and the level of similarity of demographics between pre-testing participants and sample population. | -- |
| | 5d | Questionnaire if possible, should be fully provided (in the article, or as appendices or as an online supplement). | Supplementary information |
| Sample characteristics | 6a | Describe the study population (i.e., background, locations, eligibility criteria for participant inclusion in survey, exclusion criteria). | 4 |
| | 6b | Describe the sampling techniques used (e.g., single stage or multistage sampling, simple random sampling, stratified sampling, cluster sampling, convenience sampling). Specify the locations of sample participants whenever clustered sampling was applied. | 4 |
| | 6c | Provide information on sample size, along with details of sample size calculation. | 4 |
| | 6d | Describe how representative the sample is of the study population (or target population if possible), particularly for population-based surveys. | N/A |
| Survey administration | 7a | Provide information on modes of questionnaire administration, including the type and number of contacts, the location where the survey was conducted (e.g., outpatient room or by use of online tools, such as SurveyMonkey). | |
| | 7b | Provide information of survey's time frame, such as periods of recruitment, exposure, and follow-up days. | 4 |

| | | | |
|---|---|---|---|
| | 7c | Provide information on the entry process:<br>–>For non-web-based surveys, provide approaches to minimize human error in data entry.<br>–>For web-based surveys, provide approaches to prevent “multiple participation” of participants. | Supplementary information |
| Study preparation | 8 | Describe any preparation process before conducting the survey (e.g., interviewers’ training process, advertising the survey). | 3 |
| Ethical considerations | 9a | Provide information on ethical approval for the survey if obtained, including informed consent, institutional review board [IRB] approval, Helsinki declaration, and good clinical practice [GCP] declaration (as appropriate). | 3 |
| | 9b | Provide information about survey anonymity and confidentiality and describe what mechanisms were used to protect unauthorized access. | 3, Supplementary information |
| Statistical analysis | 10a | Describe statistical methods and analytical approach. Report the statistical software that was used for data analysis. | 4 |
| | 10b | Report any modification of variables used in the analysis, along with reference (if available). | N/A |
| | 10c | Report details about how missing data was handled. Include rate of missing items, missing data mechanism (i.e., missing completely at random [MCAR], missing at random [MAR] or missing not at random [MNAR]) and methods used to deal with missing data (e.g., multiple imputation). | 4 |
| | 10d | State how non-response error was addressed. | N/A |
| | 10e | For longitudinal surveys, state how loss to follow-up was addressed. | N/A |
| | 10f | Indicate whether any methods such as weighting of items or propensity scores have been used to adjust for non-representativeness of the sample. | N/A |
| | 10g | Describe any sensitivity analysis conducted. | N/A |
| **Results** | | | |
| Respondent characteristics | 11a | Report numbers of individuals at each stage of the study. Consider using a flow diagram, if possible. | N/A |
| | 11b | Provide reasons for non-participation at each stage, if possible. | N/A |
| | 11c | Report response rate, present the definition of response rate or the formula used to calculate response rate. | Unknown due to multiple administration routes. Highlighted as limitation |
| | 11d | Provide information to define how unique visitors are determined. Report number of unique visitors along with relevant proportions (e.g., view proportion, participation proportion, completion proportion). | N/A |
| Descriptive results | 12 | Provide characteristics of study participants, as well as information on potential confounders and assessed outcomes. | 4, Supplementary materials Table 1 |

| | | | |
|---|---|---|---|
| Main findings | 13a | Give unadjusted estimates and, if applicable, confounder-adjusted estimates along with 95% confidence intervals and p-values. | Descriptive analysis only |
| | 13b | For multivariable analysis, provide information on the model building process, model fit statistics, and model assumptions (as appropriate). | N/A |
| | 13c | Provide details about any sensitivity analysis performed. If there are considerable amount of missing data, report sensitivity analyses comparing the results of complete cases with that of the imputed dataset (if possible). | N/A |
| **Discussion** | | | |
| Limitations | 14 | Discuss the limitations of the study, considering sources of potential biases and imprecisions, such as non-representativeness of sample, study design, important uncontrolled confounders. | 9 |
| Interpretations | 15 | Give a cautious overall interpretation of results, based on potential biases and imprecisions and suggest areas for future research. | 7-9 |
| Generalizability | 16 | Discuss the external validity of the results. | 9 |
| **Other sections** | | | |
| Role of funding source | 17 | State whether any funding organization has had any roles in the survey's design, implementation, and analysis. | Provided to journal at submission |
| Conflict of interest | 18 | Declare any potential conflict of interest. | Provided to journal at submission |
| Acknowledgements | 19 | Provide names of organizations/persons that are acknowledged along with their contribution to the research. | N/A |

**Survey instrument**

1. Which region do you work in?
   ☐ England
   ☐ Scotland
   ☐ Wales
   ☐ Northern Ireland
2. What age are you?
   ☐ <30 years
   ☐ 30-39 years
   ☐ 40-49 years
   ☐ 50-59 years
   ☐ 60+ years
3. What gender are you?
   ☐ Male
   ☐ Female
   ☐ Non-binary
   ☐ Prefer not to say
4. How many years of practice/experience do you have since qualifying?
   ☐ <5 years
   ☐ 5-10 years
   ☐ 10-20 years
   ☐ 20+ years
5. Please select the category which best describes your current role:
   ☐Stroke Physician
   ☐Research Nurse
   ☐ Other Physician
   ☐ Stroke Nurse Practitioner / Specialist Nurse
6. Proportion of your role spent in research:
   ☐ <10%
   ☐ 10-25%
   ☐ 25-50%
   ☐ >50%
7. Do you believe that a platform trial would be a good option for testing potential interventions for patients with stroke due to intracerebral haemorrhage?
   ☐ Strongly Agree
   ☐ Agree
   ☐ Neutral
   ☐ Disagree
   ☐ Strongly Disagree
8. Please, rate the following five information and consent-related factors affecting recruitment to ICH platform trials using the scale from strong barrier to strong facilitator.

| | strong barrier | weak barrier | neutral | weak facilitator | strong facilitator |
|---|---|---|---|---|---|
| 1. Provision of brief user-friendly information for potential participants, e.g. a video animation | | | | | |
| 2. Option to provide potential participants with tailored information on studies they are eligible for based on their care so far and management uncertainties | | | | | |
| 3. Option to share study information with potential participants or their representatives electronically in addition to traditional paper methods | | | | | |
| 4. In hyperacute settings, ability to seek verbal consent to participate before obtaining written consent | | | | | |
| 5. Option to obtain telephone or electronic consent in addition to traditional paper methods | | | | | |

9. Please, rate the following six clinical-team-related factors affecting recruitment to ICH platform trials using the scale from strong barrier to strong facilitator.

| | strong barrier | weak barrier | neutral | weak facilitator | strong facilitator |
|---|---|---|---|---|---|
| 1. Research question perceived as important | | | | | |
| 2. Research-positive culture in the clinical team | | | | | |
| 3. Clinical team with prior research experience | | | | | |
| 4. Clinician preference for specific treatments | | | | | |
| 5. Availability of designated research team | | | | | |

| |
|---|
| 6. Availability of research staff out-of-hours |

10. Are there any other modifiable facilitators which you think could aid recruitment to a future platform trial?

    Please provide details below or write "No". Do not include any identifiable information in your responses.

11. Are there any other modifiable barriers which you think might limit recruitment to a future platform trial?

    Please provide details below or write "No”. Do not include any identifiable information in your responses.

**Free text responses in time order**

| **Are there any other modifiable facilitators which you think could aid recruitment to a future platform trial?** |
|---|

Research Nurse being delegated task of taking consent and doctor not on delegation log being ethically approved to check inclusion/exclusion criteria. (If enrolment is time-dependent).

Pragmatic Design Easy Randomisation ie. Next blinded pack of drugs

Next of kin involvement Research practitioner being embedded with in the clinical team

Options of different languages available, either video or written.

Tangiable reward for recruitment even if it is just verbal recognition

Being able to gain Professional consent- e.g. from neutral ED Consultant. Pragmatic trials- not expecting extra imaging than that done clinically already.

posters for patients and relatives to see on wards

Training staff to improve understanding of the importance of the platform trial, to the point that it becomes part of their standard practice to consider patients for a platform trial, rather than viewing it as 'extra work'. Regular refresher/ reinforcement learning for staff.

The availability of Trial Nurses to facilitate recruitment by direct involvement with patients and to support existing Stroke Nurses in the Acute Setting to recruit into clinical trials

Personnel resource and infrastructure to support multi-site delivery of trial. National partnership strategy for ICH research delivery.

information (written/visual) in common languages other than English

Process needs to be as simple as possible for the sites. Minimum data collection, central follow-up

Information must be provided in aphasia friendly/easy reading format. Too often, by the time research has gone through the ethics process, the information is twice as long and half as understandable.

Development of Patient specific reminders to seek agreement to participate when a diagnosis of ICH is made

clinical training in different research studies and allowing clinicians more time for research

I think the reality of most busy clinical teams si that they are so busy firefighting that putting patients into studies plays second fiddle

Simplified PIS or other materials to support patient's and family's understanding of the trial.

Deferred consent will be essential.

Integrating NHS clinicians and nurses with research teams.

More funding for research dedicated staff and research education for clinical staff. This would provide a more positive research culture. Mandating research activities to all clinical staffs job plans.

Quick access to easy and patient friendly info about research activities and availability of research and clinical staff

more doctors/ medical staff on the delegation log and available to check eligibility/ consent patients

process to be as simple as possible everything in one place so easy to access clear signposting to suitable research options for any given person

Nhs app consent

promotion of research to be a core role of all staff members with some small funded /job planned time hence can have clinical staff with interest in research to facilitate out of hours recruitment does not have to be full time research staff being able to use potential remote verbal consent for consultee

provide research opportunities for the members of the whole MDT to improve the screening and recruitments.

A good research question

Easy randomisation processes (eg. QR code scanning as per TICH3)

Multi language option for pis/consent Wide dissemination about the trial in places patients would reach eg local media/radio

I think there needs to be equipoise amongst clinicians to engage in such trials

**Are there any other modifiable barriers which you think might limit recruitment to a future platform trial?**

Lack of staff availability.

Patient cognition lack of family support

Research burden on participants.

Consent having to be got from patient or personal representative. Trials with clinical consent, have done well in some settings. 'Blood Pressure to be less than'... criteria mRS criteria..

it needs to be straightforward, and most importantly fast, so as not to prevent established (but not necessarily evidence based) treatments from being given rapidly.

Adaptation of consenting to ensure people with communication and cognitive difficulties can participate.

caseload pressures, work place shortages of staff

Education of the public on the importance of platform trial/ clinical trials

Lack of designated/paid clinician time to perform research alongside clinical duties

Funding, multi-site partnership and leadership. Existing research silos. Portfolio income from recruitment eg. if a participant was enrolled in a platform trial then accrual based income would only be awarded once whereas in multiple trials income would be awarded for each trial - there are significant implications.

Some participants might reject any trial study resembling RECOVERY based on their personal interpretation of COVID research and perceptions of 'rushed' trial initiation

Communication with patient.

Delays in getting centres to a state of readiness

Busy clinical environements and job plans with no time for research

No. Main barriers are having appropriately interested and qualified staff available to inform, consent and enrol patients, particularly in the out-of-hours time period.

MDT awareness of trials -->Include the wider MDT in the identification of patients to support recruitment.

Can sites only host certain interventions?

availability of clinic space.

reducing red tape for ethics/approvals at individual centre R and Ds

Not having a research positive culture. Not enough funding for research staff and education.

poor perception of importance of clinical research and cultural departmental barriers against research mainly due to under-recognition of the importance of allocated research time in clinicians diary

clinician availability

information on how easy to use and accessible this structure/format could be for those less used to this study design/format

Pt information on what available on the market and consent in advance

Difficulty in getting uniformity in ICH management e.g treatments on front door, reversal of anticoagulation, which patients are referred to neurosurgeons for example. Also intensive monitoring on HASU in a trial setting may be difficult e.g. monitoring BP. Staff are usually stretched

Having easy to use EPR systems which could flag an alert to patient being suitable for a study if report states ICH/Bleed as a prompt to teams

High intensity nursing interventions (eg. 5 minute BP checks) which non-research clinical staff struggle to do or be motivated to do!

Clinician Time (clinicians require job planning to continue to contribute to research)
